# An Oxford Nanopore Technology-Based Hepatitis B Virus Sequencing Protocol Suitable For Genomic Surveillance Within Clinical Diagnostic Settings

**DOI:** 10.1101/2024.01.19.24301519

**Authors:** Derek Tshiabuila, Wonderful Choga, San E. James, Tongai Maponga, Wolfgang Preiser, Gert van Zyl, Monika Moir, Stephanie van Wyk, Jennifer Giandhari, Sureshnee Pillay, Ugochukwu J. Anyaneji, Richard J. Lessells, Yeshnee Naidoo, Tomasz Janusz Sanko, Eduan Wilkinson, Houriiyah Tegally, Cheryl Baxter, Darren P. Martin, Tulio de Oliveira

## Abstract

Chronic hepatitis B virus (HBV) infection remains a significant public health concern, particularly in Africa, where there is a substantial burden. HBV is an enveloped virus, with isolates being classified into ten phylogenetically distinct genotypes (A – J) determined based on full-genome sequence data or reverse hybridization-based diagnostic tests. In practice, limitations are noted in that diagnostic sequencing, generally using Sanger sequencing, tends to focus only on the S-gene, yielding little or no information on intra-patient HBV genetic diversity with very low-frequency variants and reverse hybridization detects only known genotype-specific mutations. To resolve these limitations, we developed an Oxford Nanopore Technology (ONT)-based HBV genotyping protocol suitable for clinical virology, yielding complete HBV genome sequences and extensive data on intra-patient HBV diversity. Specifically, the protocol involves tiling-based PCR amplification of HBV sequences, library preparation using the ONT Rapid Barcoding Kit, ONT GridION sequencing, genotyping using Genome Detective software, recombination analysis using jpHMM and RDP5 software, and drug resistance profiling using Geno2pheno software. We prove the utility of our protocol by efficiently generating and characterizing high-quality near full-length HBV genomes from 148 left-over diagnostic Hepatitis B patient samples obtained in the Western Cape province of South Africa, providing valuable insights into the genetic diversity and epidemiology of HBV in this region of the world.

## Introduction

Chronic hepatitis B virus (HBV) infection is a significant global health concern, leading to severe outcomes such as hepatocellular carcinoma (HCC) and liver cirrhosis. It has a high intermediate prevalence in sub-Saharan Africa, where approximately 6.1% of adults are chronically infected, contributing to a substantial portion of the 256 million global cases [1]. The African region has a highly endemic nature of chronic hepatitis B (CHB), accounting for a quarter of cases and witnessing around 1.5 million new infections annually. In 2019, the global HBV prevalence was estimated at 4.1%, with the Western Pacific region having the highest prevalence (7.1%) and the European region the lowest (1.1%) [2, 3].

HBV, a member of the *Hepadnaviridae* family, possesses a compact circular genome of approximately 3.2 kilobases (kb) [4]. It comprises four genes—HBx (X), Core (Pre-C/C), Surface (S), and Polymerase (P)—with seven overlapping reading frames. The S protein, containing the “a”, determinant region within the major hydrophilic region (MHR), is crucial as it serves as the target for anti-HBs antibodies. Substitutions in and around this determinant region are associated with immune escape and vaccine failure [5].

To date, ten HBV genotypes (A-J) with an intergroup divergence of at least 8% at the whole genome level were identified [6]. The most prevalent genotypes globally are C (26%), D (22%), E (18%), A (17%), and B (14%) [7]. In Africa, predominant circulating genotypes include A, D, and E, with sub-genotypes A1 and D3 predominating in southern Africa. Specifically, sub-genotype A1 remains most prevalent (accounting for 70 – 90% of infections; in South African populations and has been linked to severe liver disease and rapid progression to HCC [8, 9].

While whole-genome sequencing-based genome surveillance is becoming a key tool for understanding the distribution, infection prevalence, and genetic diversity of HBV and other viral pathogens [10–12], it has yet to be fully leveraged in clinical diagnostic settings. In such settings, serological tests are used to detect HBV surface antigen (HBsAg) in serum or plasma. However, a major concern when using serological screening tests is that these tests must possess a high degree of sensitivity and an acceptable level of specificity to reduce false results [13, 14]. For more detailed characterization of HBV in patient samples, Sanger sequencing of complete or partial HBV genomes is considered the gold standard. However, partial genome sequences can be somewhat misleading when characterizing mixed-genotype (i.e. recombinant) HBV genomes, and Sanger sequencing yields little or no information on intra-patient HBV genetic diversity which may result in drug resistance, assist in vaccine development, and help in understanding pathogenesis and transmission patterns [15].

High-throughput sequencing (HTS) techniques are powerful tools that, in addition to diagnosing CHB and genotyping of HBV, enable the detection of viral drug resistance mutations, the identification of recombinant HBV genomes (including those with mixed genotypes), and the assessment of intra-patient HBV diversity [16]. Among numerous other HBV-focused applications, HTS and downstream analyses have previously been used to track the diversity of HBV populations within individual CHB patients [17], identify the prevalence of drug-resistance mutations in large patient cohorts [15, 18–20], and sequence complete HBV genomes [10, 11, 21, 22].

Two major challenges associated with HTS workflows employed to generate viral genome sequence data are 1) the efficient and accurate barcoding of samples needed for multiplexed sequencing where multiple patient samples are simultaneously sequenced in a single run and 2) the accurate reassembly of sub-genome-length sequence reads into complete genomes. Third-generation HTS technologies, such as the Oxford Nanopore Technology (ONT) have largely overcome these limitations. Barcoding kits allow for cost-effective and efficient sequencing by pooling and running multiple libraries on a single flow cell. Different types of barcoding kits are available, including ligation-based, PCR-based, and rapid transpose-based kits, each with its own advantages and input requirements (https://community.nanoporetech.com/docs). A study by McNaughton et al. describes advancements in a sequencing protocol utilizing isothermal rolling-circle amplification and ligation-based barcoding kits [23]. However, the challenges associated with rolling circle amplification include the generation of concatemers of the HBV genome instead of a single genome. The ONT rapid barcoding Kit does not require individual sample washes and allows samples to be processed uniformly without quantification or normalization [24]. The sample runtime on ONT platforms, per 96 samples, is almost half that of Illumina (14 hours compared to 26 hours) mainly due to real-time data analysis with ONT. The rapid barcoding library preparation utilized by ONT also requires fewer reagents, as everything is provided within the kit, and is thus cheaper than Illumina sequencing [25]. Further, the sequence reads generated using ONT are substantially longer than those generated by Illumina, a factor that vastly simplifies the assembly of whole genome sequences. However, ONT still exhibits a higher error rate than Illumina sequencing although this has improved with newer chemistry and the use of post-sequencing software such as Nanopolish.

Here we describe an optimized ONT-based HBV whole genome sequencing protocol with associated downstream computational analyses. Using 148 samples from leftover diagnostic samples sent for HBV DNA load testing of South African patients with hepatitis B, we demonstrate that the protocol is applicable in clinical settings for both the accurate recombination-aware genotyping of patient samples and the detection of drug-resistance mutations.

## Results

### Sequencing of samples

To evaluate the applicability of our ONT-based protocol in a clinical setting, we applied it to 148 HBV-positive samples collected between September 19, 2022, and November 5, 2023, in Western Cape, South Africa. The average age of the patient cohort was 40 years (range: 0 to 69), with a gender distribution of 37.2% females and 60.8% males, and three samples had no information for gender. The overall median viral load (VL) was 16,368 IU/ml (IQR: 1,452 - 1,445,182) (**Table 1**). Of the 148 samples sequenced, 146 HBV genomes (98.6%) were obtained. Samples with missing age, gender, and viral load information were excluded from the analyses and the 138 sequences with available metadata were used for subsequent analyses.

**Table 1:** Demographic characteristics for the 148 patient samples.

| Characteristic | Total N=148 (%) |
| --- | --- |
| **Gender** |  |
| Male | 90 (60.8) |
| Female | 55 (37.2) |
| Unknown/Other | 3 (2.0) |
| **Age** |  |
| <18 | 3 (2.03) |
| 18 - 24 | 6 (4.05) |
| 25 - 34 | 34 (22.97) |
| 35 - 44 | 45 (30.41) |
| >=45 | 56 (37.84) |
| Missing | 4 (2.7) |
| **HBV viral load (IU/ml)** |  |
| <2000 | 43 (29.05) |
| 2000 - 20 000 | 52 (35.14) |
| >20 000 | 52 (35.14) |
| Missing | 1 (0.67) |

The 138 complete HBV genomes with available metadata had a median sequencing depth of 2344 (IQR 584-13 497) and median genome coverage of 98.33% (IQR 92%-100%). Of the 138 sequences, 123 had a uniform coverage of greater than 80%. The GD Hepatitis B phylogenetic typing tool classified 114 of the 138 near-complete genomes and determined that the majority (n=103, 74.6%) were genotype “A,” followed by “D” (n=9, 6.5%) and “E” (n=2, 1.4%). The 24 genomes (17.5%) that GD could not assign to a genotype level had a degree of genome coverage that was significantly lower than that of the genomes that could be assigned (median coverage of 92.74% vs 98.68%; p-value 0.003472; Wilcoxon rank sum test). Further analysis of genome coverage revealed a statistically significant difference in coverage between genotypes A and D (p-value 0.00884) (**Figure 1A**). Although viral load did not significantly affect the genome coverage, a weak linear association (R^2^ = 0.099; Pearson linear correlation) was observed between high viral load and genome coverage (**Figure 1B**).

**Figure 1:**
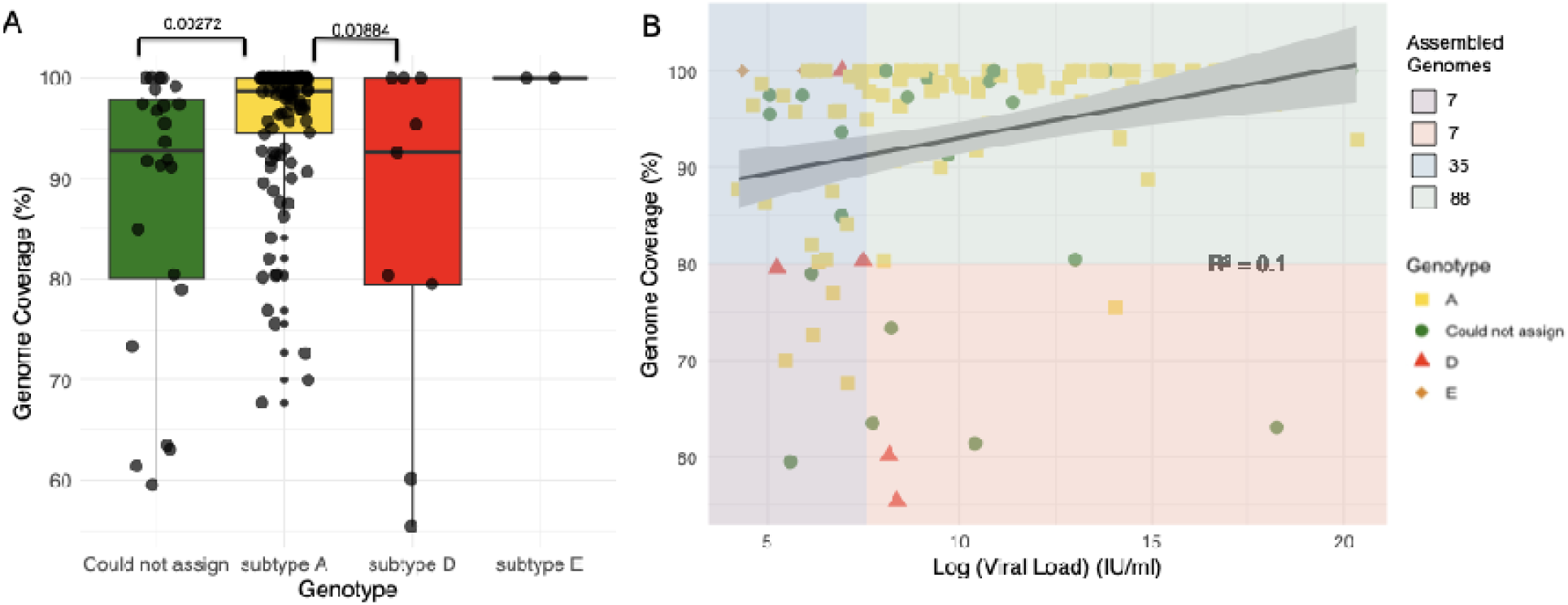
(A) Comparison of genome coverage for different detected HBV genotypes. The boxes indicate the lower quartile, median, and upper quartile, and the minimum and maximum values by the whiskers. Significant differences between genome coverage (paired Wilcoxon test) values between each genotype are denoted above the box and whisker plots. **(B) Scatter plot fo log viral load against the genome coverage for detected HBV genotypes.** A total of 123 genomes with >80% coverage were produced (35 with a log viral load of <7.6 IU/ml and 88 with a log viral load of >7.6 IU/ml). HBV genotypes are represented by different colors and shapes. The different background color shades represent different quality control groups. The blue and purple shades represent ≤7.6 IU/ml log viral load and coverage of ≥80% and <80%, respectively. The green and pink shades represent a log viral load of >7.6 IU/ml and coverage of ≥80% and <80%, respectively.

### Recombination Detection and Breakpoint Identification

RDP5.46 and jpHMM were used to identify recombinant sequences and infer breakpoints. jpHMM maps were generated for the 24 “unassigned” study sequences to infer evidence of inter-genotype recombination. Of the 24 sequences, traces of recombination were detected in 16, and the remaining eight sequences were assigned to genotypes by jpHMM (6 D, 1 A, 1 G) (**Figure 2**). jpHMM classified twelve sequences as A/D recombinants (i.e. the parents of the recombinants belonged to genotypes A and D) (**Figure 3**), three as A/E recombinants, and one as an A/D/G recombinant (**Figure 4**). Bootscan plots and genome coverage maps were obtained from the GD Hepatitis B phylogenetic typing tool to assess the accuracy of the non-A/D recombination classifications (Figure 4).

**Figure 2:**
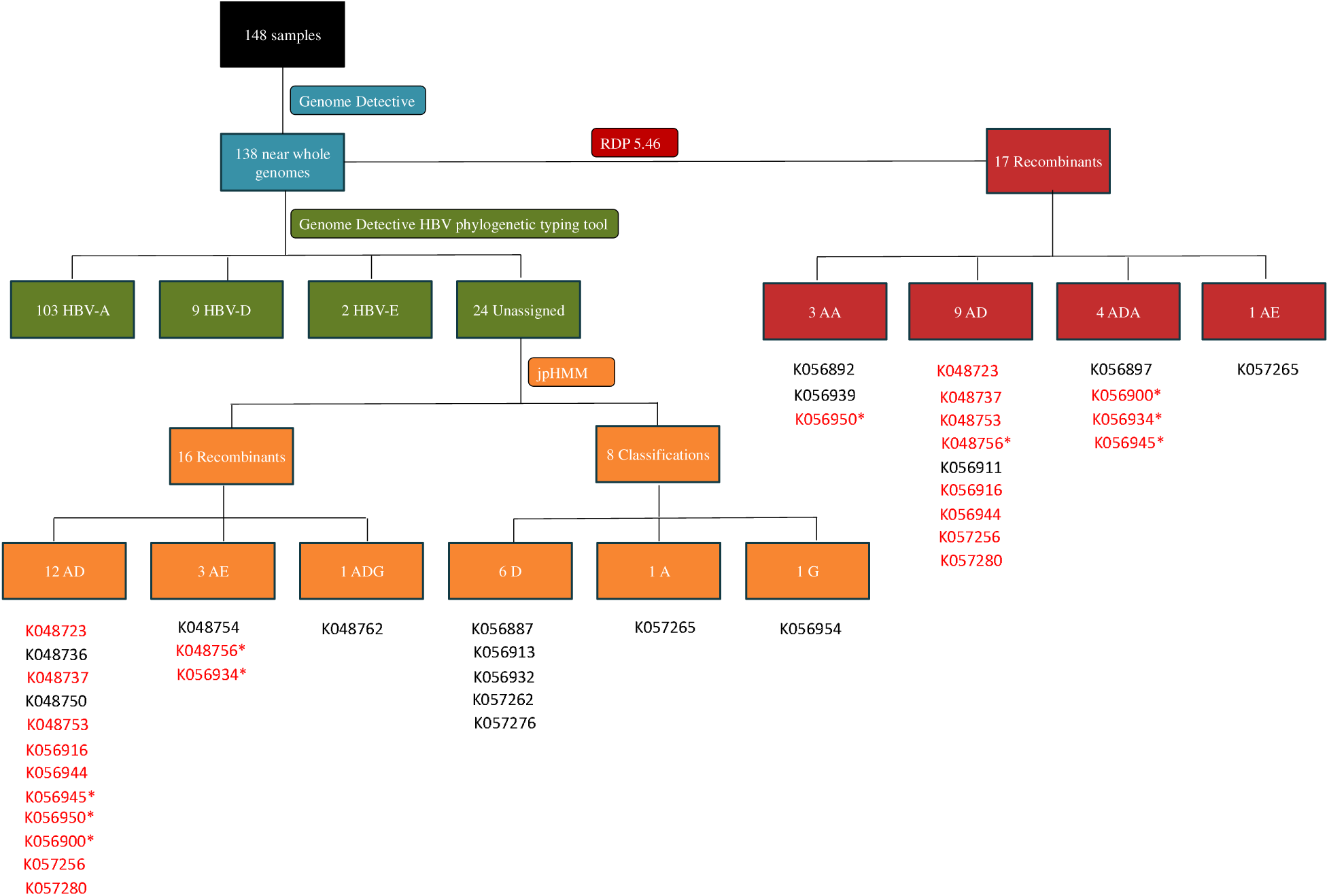
Flowchart showing the identification of recombinants using the jpHMM HBV-tool and RDP5.46. The blue section shows the number of genomes produced by Genome Detective, the green section shows genotype variation as assessed by the Genome Detective Hepatitis B phylogenetic typing tool; the orange section highlights the jpHMM recombination classifications of the unclassified sequences, and the red section shows the RDP5.46 recombination analysis. Sample IDs are shown below the flowchart sections and IDs that are shown in red highlight recombinants that both jpHMM and RDP5.46 detected. Sample IDs marked with a “*” were classified as different recombinants by jpHMM and RDP5.46.

**Figure 3:**
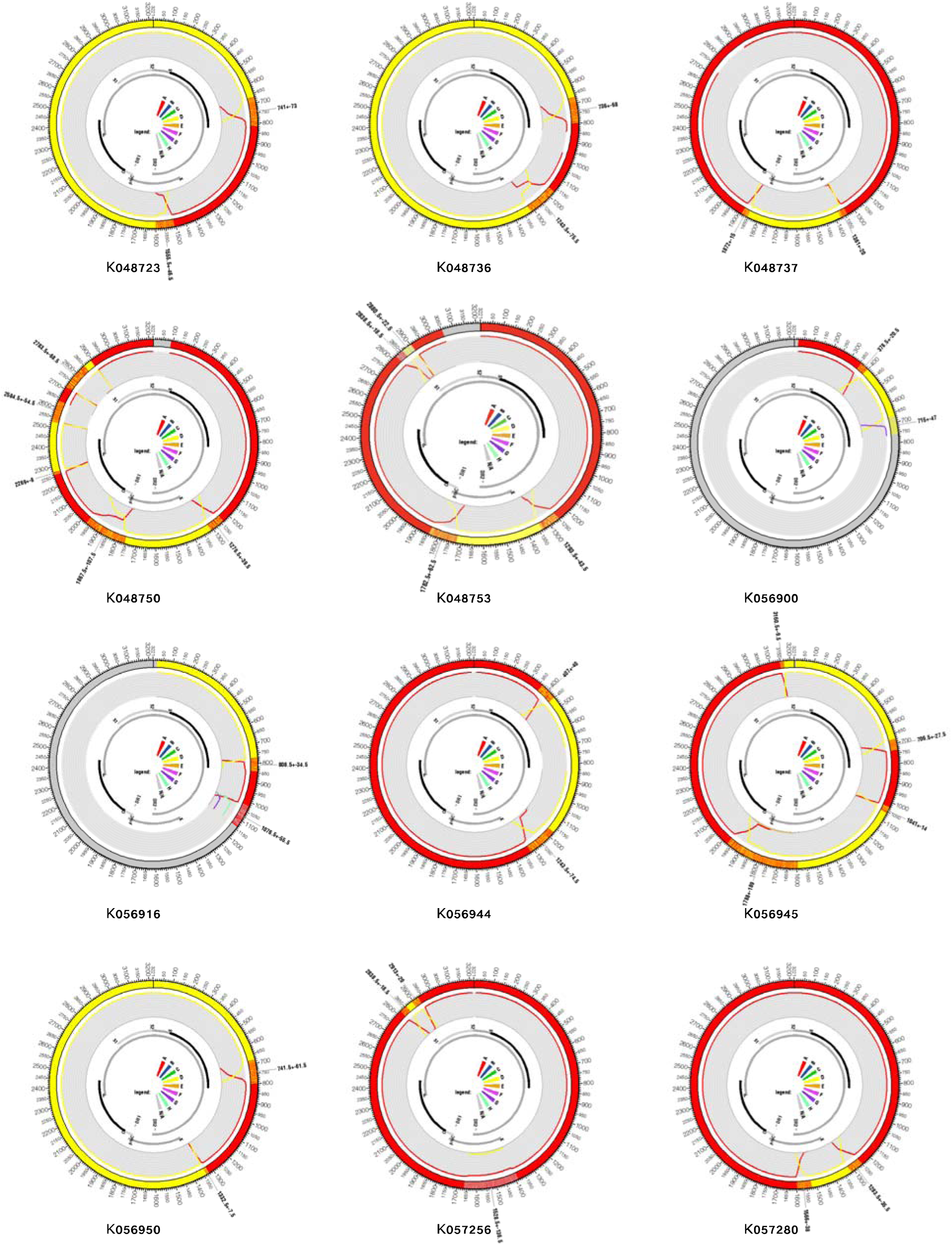
jpHMM genome maps for A/D recombinant viruses. The query isolates identifier names are listed below each jumping profile map. Genome maps presented here were created using the software package Circos [26]. The colored shadings represent different HBV genotypes (red = A, yellow = D, grey = unknown). Regions of orange shading represent recombination breakpoint intervals, e.g. region 405 ± 40 (outer ring). All sequence position numbers are given relative to the HBV reference genome AM282986. Positions of genes in the genome are marked with grey and black bars (inner ring). The color legend is in the middle, and the “NA” denotes “not assigned”.

**Figure 4:**
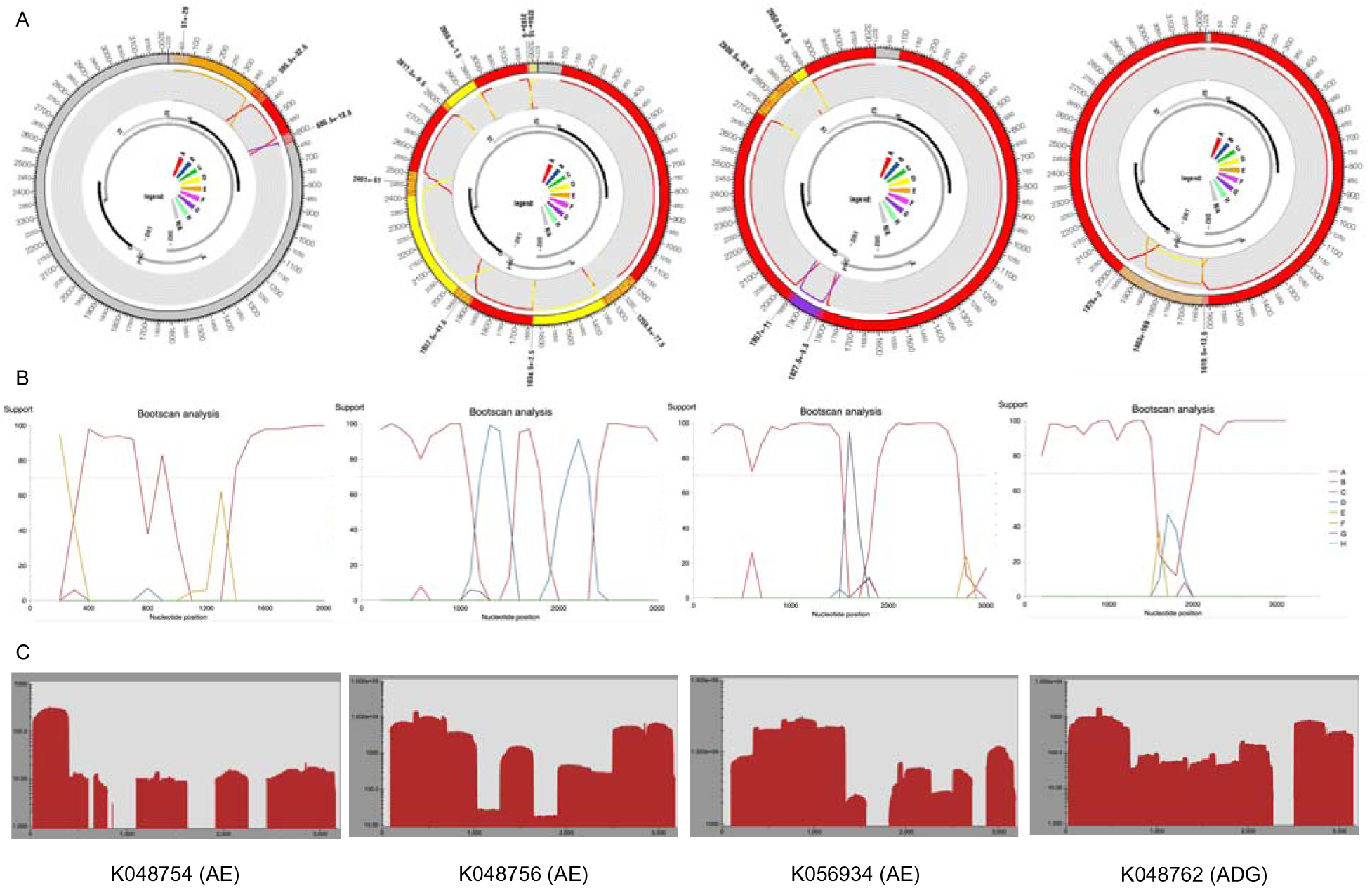
(A) jpHMM genome maps and (B) Genome Detective bootscan plots for non-A/D recombinant viruses. Bootscan analysis was performed with a window size of 400 and a step size of 100. **(C) Genome coverage maps highlighting HBV sequencing depth.** The query isolates are listed below each genome coverage map.

Further assessment of recombination using RDP5.46 but without assuming only recombination between subtypes (i.e. also accounting for intra-genotype recombination) confirmed that 12 of the 16 recombinants identified by jpHMM were actual recombinants (Figure 2). P-values are listed in the supplementary file, Table S2. Of the twelve sequences classified as A/D recombinants by jpHMM, RDP5.46 confirmed seven as A/D recombinants, with the remaining five classified as two A/D/A recombinants, one an A/A recombinant, and two as non-recombinants. Two A/E recombinants were classified as A/D and A/D/A recombinants by RDP5.46 (Figure 2). Recombination events were distributed variably, with recombination breakpoints concentrated towards the end of the P gene, within the X and pre-C regions, and at the start of the C region (**Figure 5B**). The recombination region count matrix illustrated heightened susceptibility for recombinational transfers of specific genomic regions, particularly the end of the pol region, the X region, the pre-C region, and the C region (**Figure 5A**).

**Figure 5:**
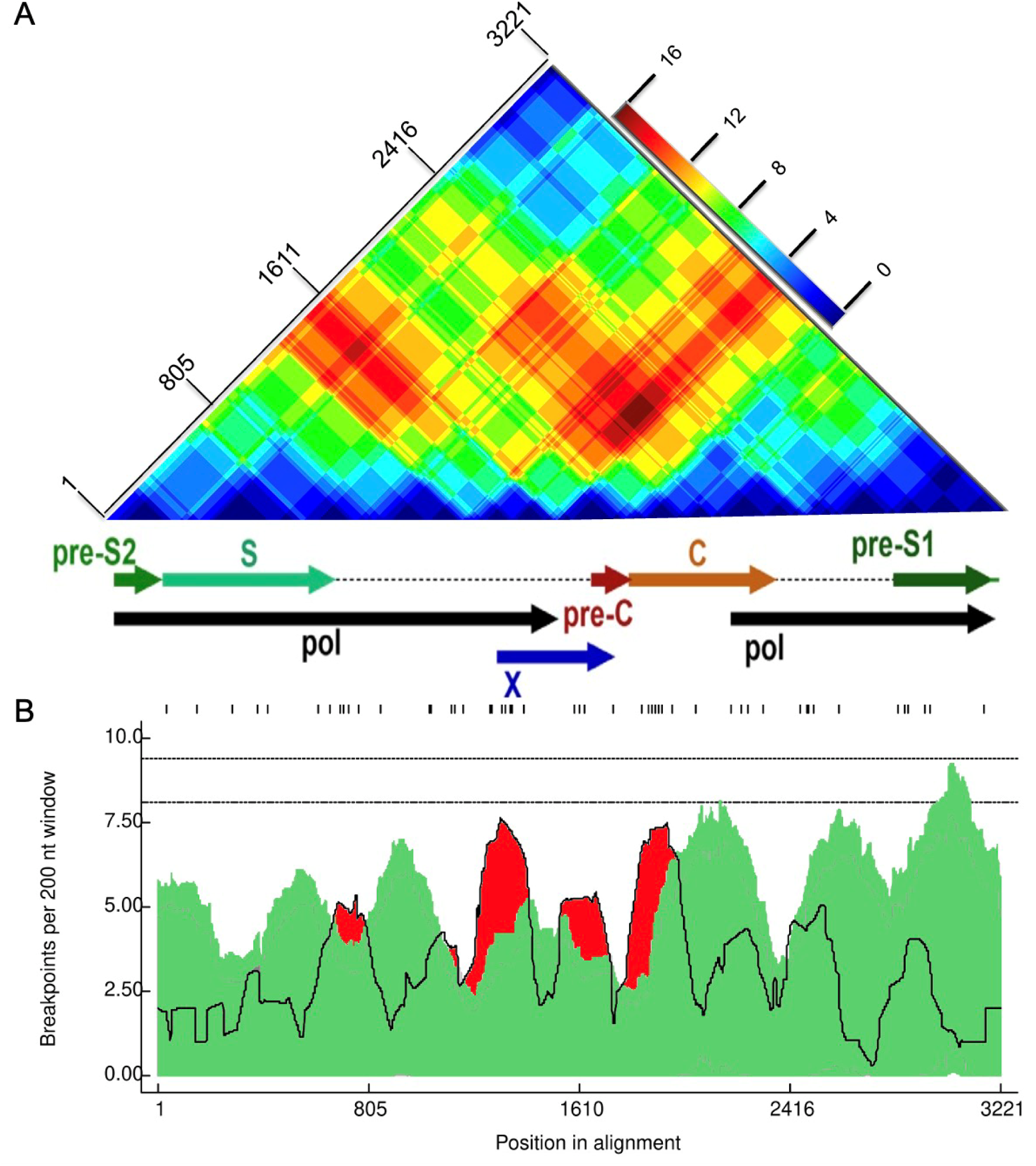
Recombination region and breakpoint distributions (A) recombinant region count matrix highlighting areas of the genome that are most and least commonly transferred during recombination events. Unique recombination events were mapped onto a region count matrix based on determined breakpoint positions. Each cell in the matrix represents a pair of genome sites with the colors (heat) of cells indicating the number of times recombination events separated the represented pairs of sites. **(B) Breakpoint distribution across HBV genomes.** All detectable breakpoint positions are represented as black lines above the graph. The green areas show the 99% confidence interval for breakpoint clustering under random recombination. The upper dotted line represents the global 99% confidence interval for a breakpoint clustering under random recombination and the lower dotted line is the global 95% confidence interval for breakpoint clustering under random recombination. The black line represents the number of breakpoints within a 200-nucleotide window moved along the genome. Areas in red where the black line emerges above the green area are considered recombination warm spots and those that traverse the dotted global 95% confidence interval line are considered statistically supported recombination hotspots. An HBV gene map is plotted between the figures.

Throughout the study period (September 2022 - November 2023), subtype A (n = 103) was the most prevalent HBV genotype, while subtype D (n = 9) only occurred sporadically in September 2022 and February, May, June, and July 2023. Subtype E (n = 2) was identified only in October 2022. Recombinant genomes were sampled throughout the study period (**Figure 6**). This temporal variation in subtype prevalence underscores the dynamic nature of HBV transmission and the importance of continuous surveillance.

**Figure 6:**
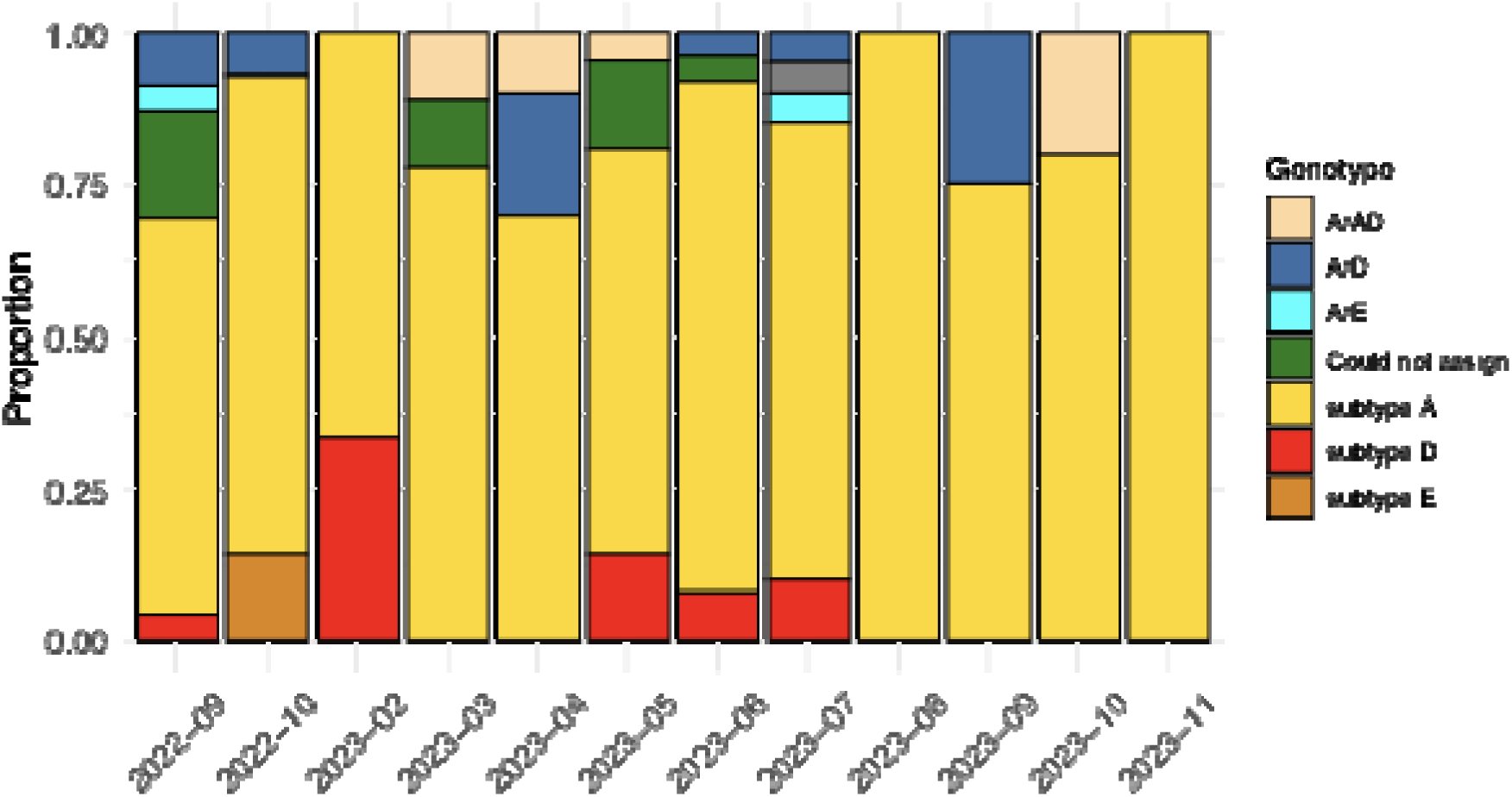
Frequency and distribution of HBV genotypes circulating in South Africa between September 2022 and November 2023. Genotyping was performed using the GD HBV phylogenetic typing tool and recombination analysis was performed using RDP5.46.

### Drug resistance profiling

Mutations related to HBsAg escape and drug-resistance of RT/HBsAg overlapping region were determined using Geno2pheno-HBV. Among the 138 sequences, 18 (13.08%) were likely resistant to lamivudine and telbivudine. Fifteen (10.87%) were resistant to entecavir, and three (2.17%) were resistant to adefovir. All sequences were, however, likely susceptible to th tenofovir drug class (**Figure 7A**). 204V and 180M are the most prevalent drug-resistance mutations (**Table 2**). The most common vaccine escape HBsAg mutation is 100C, which i responsible for HBV detection failure [27], with a frequency of five. The second most prevalent HBsAg mutation is 120T, which is responsible for vaccine, immunotherapy, and diagnosti detection failure, with a frequency of 2. This is followed by mutations 128V, 109I, and 101H, each with a frequency of 1, which are responsible for vaccine escape (**Figure 7B**) [27].

**Figure 7:**
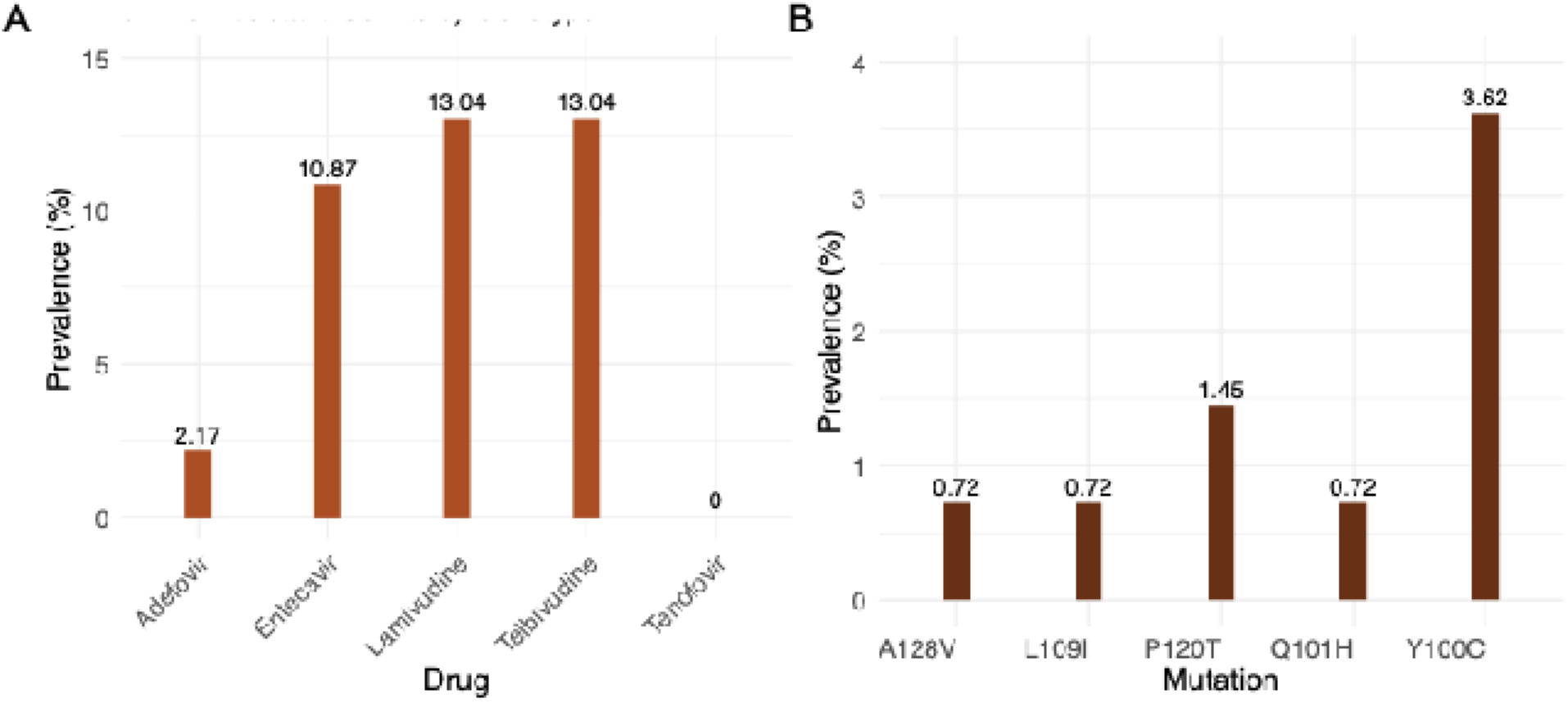
(A) Prevalence of predicted drug resistance based on mutation patterns in the RT/HBsAg overlapping region for HBV genomes sampled in South Africa between September 2022 and November 2023. (B) Prevalence of HBsAg vaccine escape mutations in the RT/HBsAg overlapping region for these same genomes.

**Table 2:** Prevalence of drug-resistant mutations in the RT/HBsAg overlapping region.

| Mutation | Adefovir | Entecavir | Lamuvudine | Telbivudine |
| --- | --- | --- | --- | --- |
| 180M | 0 | 4 | 4 | 0 |
| 204V | 0 | 5 | 5 | 5 |
| 181T | 1 | 0 | 0 | 0 |
| 202K | 0 | 2 | 0 | 0 |
| 250S | 0 | 1 | 0 | 0 |
| 250N | 0 | 1 | 0 | 0 |

## Discussion

In this study, we demonstrate that ONT sequencing, utilizing the Oxford Nanopore Rapid Barcoding Kit, enables the rapid and simple generation of full-length HBV genomes. Additionally, we illustrate the utility of the rich sequencing data generated by this approach in the recombination-aware genotyping of HBV genomes and the detection of mutations associated with drug resistance and vaccine/immunotherapeutic resistance.

Sequences produced using our protocol resulted in 123 genomes with uniform coverage of greater than 80% and a sequencing depth of approximately 2343.86. Sanger sequencing is considered the gold-standard DNA sequencing method and has been used to classify HBV into its ten genotypes (A-J). It is, however, often restricted to analyzing specific genes and is rarely used for the analysis of intra-patient genetic diversity [15]. Illumina deep sequencing, while effective for genotyping and characterizing genetic diversity [28], suffers from limitations such as the inability to sequence long DNA stretches, biases introduced during amplification steps, and challenges in generating sufficient overlap between DNA fragments [29]. In contrast, we opted for ONT, which overcomes these limitations with a shorter hands-on preparation time, by providing long-read sequencing, eliminating the need for complex library preparation processes, and reducing the risk of biases associated with amplification steps in Illumina sequencing. As the ONT rapid barcoding library preparation requires fewer reagents than the Illumina sequencing protocol, ONT sequencing was also much cheaper (Illumina cost per sample is ~150–250 USD whilst the ONT cost per sample is ~10–40 USD) [25]. The protocol we optimized offers a fast approach to generating HBV whole genomes as Illumina sequencing runtimes (26hrs per 96 samples) are almost double the runtimes required for ONT (14hrs) mainly due to the ability of real-time data analysis with ONT [25].

An important factor associated with the successful amplification of a virus is related to the viral load present in the sample: the higher the viral load in the sample, the higher the yield of amplified products to be sequenced and the easier it is to assemble a reliable complete genome [30]. Full-length and sub-genomic approaches have been used for ONT sequencing of HBV. However, these methods only worked well with high HBV viral loads, with one study having a raw read error rate of ~12% and the other unable to definitively confirm putative minority variants detected in the MinION reads [31, 32]. Another study by McNaughton et al. developed a sequencing protocol utilizing rolling circle amplification coupled with ONT ligation-based barcoding kits that improved the accuracy of HBV nanopore sequencing for use in research and clinical applications [23]. Our rapid, chemistry-based, barcoding kit, sequencing protocol produced complete genomes at low (<2000 IU/ml), medium (2000 - 20 000 IU/ml), and high (>20 000 IU/ml) viral loads and allowed for the identification of various HBV genotypes; including genotypes A, D, and E.

Genome characterization of a virus can be important for clinical diagnostics as, beyond identifying the infecting agent, it can reveal clinically relevant genetic variations [33]. HBV-A was the most prevalent genotype among our study cohort, which is consistent with this genotype’s high prevalence in sub-Saharan Africa [34]. Recombination analysis also revealed complex viral replication/recombination dynamics, with 16 identified recombinants, primarily between genotypes A and D (A/D). This was to be expected due to the high prevalence of genotypes A and D in Southern Africa [8, 9]. While RDP5.46 analysis confirmed eight of these recombinants, discrepancies were noted, highlighting the challenges in precisely classifying recombinant strains. Although recombination can be easily detected using phylogenetic trees and recombination software, it is much more difficult to determine whether detected recombinants exist or whether they are detection artifacts arising from 1) primer jumping during PCR of samples that are either from mixed infections or have been accidentally cross-contaminated, 2) “backfilling” of failed amplicons with contaminating sequence reads or 3) incorrectly assembled genomes where reads from multiple different genetically distinct viruses get assembled into a single genome. In general, the only way to confirm recombinants’ existence is to independently amplify and resequence samples or detect multiple genomes in different patients with identical recombination patterns (i.e., identify circulating recombinants). Of the 16 recombinants detected here, only three were identified by RDP5.46 as circulating recombinants. Therefore, until the other 13 are independently amplified and sequenced, it cannot be definitively stated that they are not simply either sequence amplification or sequence assembly artifacts. Furthermore, carry-over contamination could have arisen, particularly from highly viraemic samples, from the instruments during HBV viral load processing.

Nevertheless, the fact that HBV recombinants have been so widely and increasingly detected suggests that the recombinants detected here are likely accurate. Genotype B/E recombinants identified in Eritria are the most common in Africa [35]. Genotype D/E recombinants are also common, having been identified in eight countries, namely Kenya, Niger, Egypt, Ghana, Libya, Mali, Eritria, and Uganda [35], followed by genotype A/E, identified in five countries, namely Uganda, Eretria, Ghana, Niger, and Mozambique [35, 36], and genotype A/D, reported in Egypt, Eretria, and Uganda [35].

Also, just as we have found here, others have detected heightened levels of recombination within the C region, pre-C, P, and X genes [37]. These regions were also frequently transferred during recombination events and, as a result, may impact the diagnosis and treatment of HBV since coinfection and viral recombination can trigger greater virulence and result in a worsened patient clinical status [30].

The Geno2pheno-HBV tool is reliable for HBsAg vaccine escape and drug-resistant mutation analysis for HBV sequences [38, 39]. Here, it was used to analyze the drug-resistant profiles for the HBV genomes. The HBsAg vaccine escape mutations in this study are mutations associated with vaccine or diagnostic escape. Mutations, such as the triple mutation 173L + 180M + 204I/V, and 133L/T, found in the Pol gene, have been identified as major vaccine escape and diagnostic mutations. An HBV study in Bangladesh identified HBsAg mutant 128V as their most common mutant [27] while mutant 100C was the most prevalent in our study. This may be due to the difference in genotype prevalence between the two study populations as HBV-C was the most prevalent genotype in Bangladesh whilst genotype A is the most prevalent among our study population. One of the most prevalent resistance-associated mutations for HBV is 204V/I for both treatment-experienced and treatment individuals. This mutation can either occur alone or can occur in combination with other mutations such as 80I/V, 173L, 180M, 181S, 184S, 200V, and/or 202S [40]. Our study also notes a high prevalence of 204V/I in combination with other drug-resistance mutations.

First-line treatment for CHB includes PEGylated interferon and nucleoside/nucleotide analogs such as Tenofovir, Entecavir, and Lamivudine [9]. In South Africa, over 1.9 million people are chronically infected with HBV and the most used first-line treatment is Tenofovir in the form of tenofovir disoproxil fumarate (TDF) [9]. In this study, we note that all sequences were susceptible to Tenofovir. TDF treatment is long-term and is frequently given indefinitely due to the risk of infections reactivating when therapy is terminated [41]. A CHB functional cure, characterized by loss of HBsAg and reduced risk of HCC, can be achieved by treatment regimens including TDF [9]. However, such treatments do not eliminate the covalently closed circular DNA, and treatment termination typically results in relapse [9]. Resistance to TDF has been noted in patients harboring mutations; 80M, 180M, 184A/L, 153Q, 191I, 200V, 204V/I, 221Y, and 223A, [42]. The absence of drug-resistance mutations in genotype E genomes and their presence in genotype A and D genomes highlight the importance of monitoring drug resistance as treatment with TDF may not be successful for these patients.

A major limitation of the current study is that HBV samples were only sequenced using the proposed protocol. The sequencing data was not compared to results generated using Illumina sequencing protocols. Moreover, efforts have been invested in optimizing the analysis pipeline to enhance accessibility and enable clinical laboratory staff to execute the entire process from start to end seamlessly. This initiative seeks to empower non-specialist bioinformaticians and streamline the workflow, making genomic analysis more user-friendly and ensuring that valuable insights can be obtained without the dependency on specialized expertise. The aim is to make the application of these advanced sequencing technologies accessible to all, allowing broader adoption and application in clinical settings.

While short-read sequencing is considered the gold standard for sequencing of viral genomes [25], despite ONT having a slightly higher error rate, ONT appears to generate high-quality data at a very affordable cost. Therefore, ONT-based sequencing is presently the most cost-effective, high-throughput sequencing technology, especially well-suited for countries with limited resources for monitoring shifting viral demographics and tracking the prevalence and spread of drug resistance and vaccine evasion mutations.

The high value of genome surveillance was excellently demonstrated during the COVID-19 pandemic. Although whole genome sequencing was primarily used to monitor virus evolution and was only carried out on ~2.12% of all diagnosed COVID-19 cases (https://data.who.int/dashboards/covid19/cases?n=c, https://gisaid.org/), the prospect of agnostically diagnosing any viral pathogens (whether known or unknown) through HTS-based metagenomic sequencing is likely to be realized within the coming decade. Diagnosis augmenting protocols that are applicable in a clinical virology setting, such as the one we present here, are an important first step towards this objective.

## Materials and methods

### Study design

The 148 patient samples analyzed here in a cross-sectional study aimed at validating our ONT-based sequencing protocol and analysis pipeline were anonymized residual diagnostic plasma/serum specimens sent for HBV DNA load measurement to the National Health Laboratory Service (NHLS) based at the Division of Medical Virology, Tygerberg Hospital, South Africa. All the samples were confirmed HBV DNA-positive. The Health Research Ethics Committee (HREC) approved the study at Stellenbosch University HREC: N22/08/089.

### HBV DNA extraction

DNA was extracted from 1 ml of patient serum using the Qiagen DNA extraction kit, following the manufacturer’s instructions (QIAGEN, Hilden, Germany). Extracted DNA was eluted in a volume of 50 μl. The extracts were stored at −80 °C until use.

### Primer design

Primers were designed using Primal scheme (https://primalscheme.com) using the HBV sequences of genotypes A-J obtained from GenBank (https://www.ncbi.nlm.nih.gov/labs/virus/vssi/#/). These primers were designed to target the full length of the HBV genome (**Table 3**) and were used to obtain tiling Polymerase Chain Reaction (PCR) products for ONT library preparation.

**Table 3:**
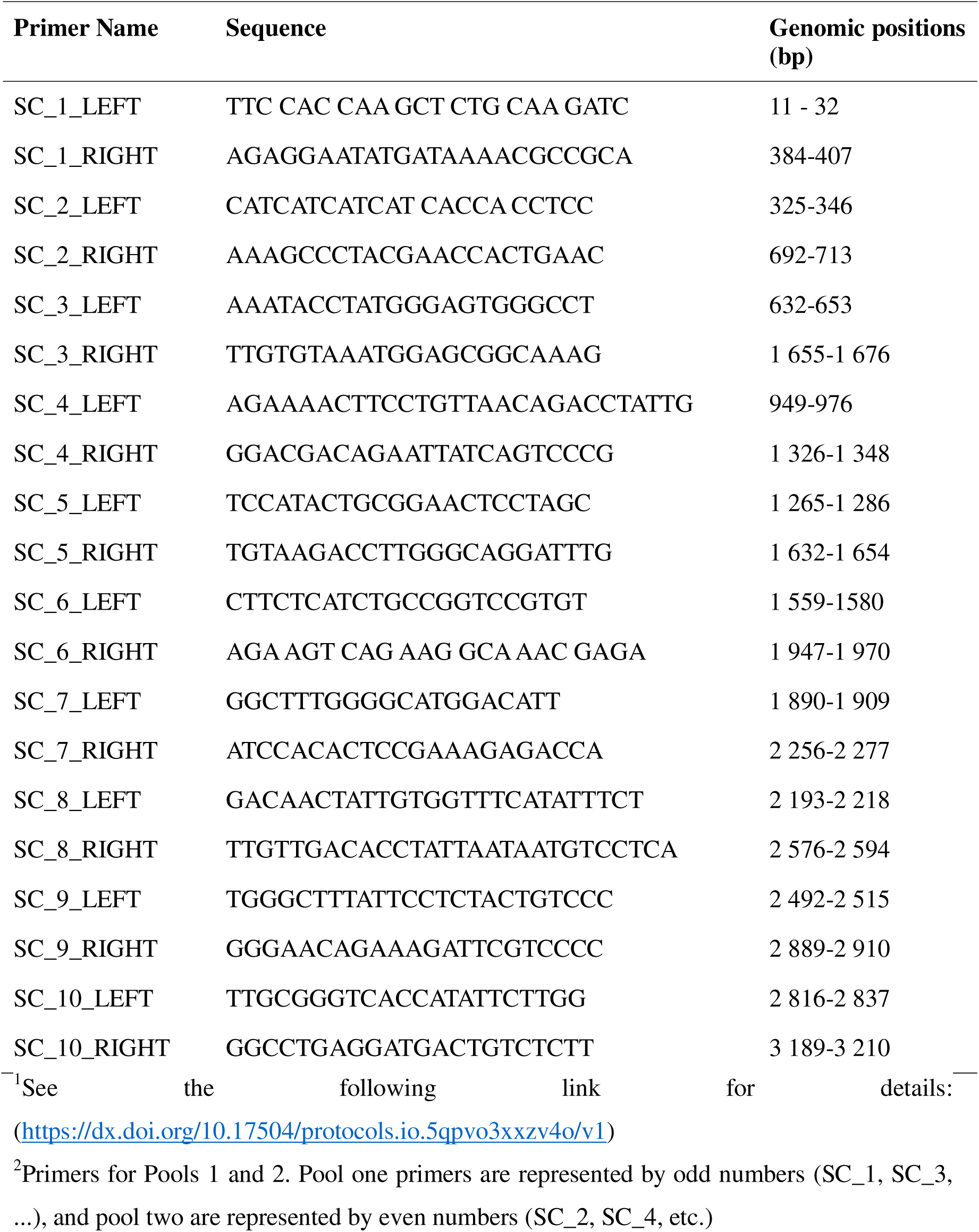
The primer sequences and genomic targets used for the tiling PCR and library preparation for ONT sequencing^1,2^.

### Tiling-based Polymerase Chain Reaction

For HBV whole-genome amplification using a multiplex PCR approach, we designed primers using Primal Scheme that would generate 1200 base pair (bp) amplicons with 70 bp overlaps, spanning the 3200 HBV genome. Since HBV has a relaxed circular DNA genome, we modified the ARTIC SARS-CoV-2 amplification protocol with proven applicability in clinical settings [43] by excluding the synthesis of complementary DNA (cDNA). Instead, we prepared master mixes using two pools of HBV primers. Randomly generated PCR products were quantified using the Qubit double-strand DNA (dsDNA) High Sensitivity assay kit on a Qubit 4.0 instrument (Life Technologies). Amplicons were purified using 1x AMPure XP beads from Beckman Coulter and quantified using the Qubit dsDNA HS assay kit from ThermoFisher. DNA library preparation was carried out using the SQK-RBK110.96 rapid barcoding kit from ONT. The resulting sequencing libraries were loaded onto an R9.4 flow cell from ONT.

### Raw-read assessment and genotyping

Whole-genome sequencing of samples was performed using the GridION platform (ONT, Oxford, UK) using the primers listed in **Table 1**, and the samples were sequenced in three runs. Subsequently, raw sequencing files underwent base calling using Guppy v3.4.5, and barcode demultiplexing was performed using qcat (demultiplexing tool) incorporated in MinKNOW Release (version 22.05.12). *De novo* genome assembly was carried out using Genome Detective (GD) 1.132/1.133 (https://www.genomedetective.com/ last accessed 03 August 2023). Briefly, GD utilizes DIAMOND to identify and classify potential viral reads within broad taxonomic units (20). The viral subset of the SwissProt UniRef protein database was used for this classification. Subsequently, reads were assigned to candidate reference sequences using the National Center for Biotechnology Information (NCBI) nucleotide database to implement a Basic Local Alignment Search Tool analyses (BLAST) and aligned using AGA (Annotated Genome Aligner) and MAFFT (v7.490) [44]. The resulting contigs and consensus sequences were then exported in FASTA file format. Raw read files were deposited in the NCBI SRA database (**Table S1**).

### Recombination analysis

All sequences in the study were assessed for inter-genotype recombination using the Jumping profile Hidden Markov Model (jpHMM) HBV tool, available at http://jphmm.gobics.de/submission_hbv, and intra- and inter-genotype recombination using recombination detection program (RDP) version 5.46 [45]. The jpHMM-HBV online tool was used to plot and visualize inter-genotype recombination breakpoints. A full exploratory automated scan for recombination with RDP5.46 was performed with the RDP [46], GENECONV [47], and MaxChi [48] methods as “primary scanning methods” to detect recombination signals and the Bootscan [49], Chimaera [50], SiScan [51], and 3Seq [52] methods to verify the signals (secondary scanning methods). The RDP5.46 general setting for sequence type was set as circular, the bootscan window size was set to 500 bp with a step size of 20 bp, and the SiScan window size was set to 200 bp with a step size of 20 bp. The remaining analysis options were kept at their default values. To ensure reliability, the HBV sequences identified as potential recombinants by RDP5.46 were only considered to be recombinant when the recombination signal was supported by at least four methods with *P*-values of ≤0.05 after Bonferroni correction for multiple comparisons [53–55].

### Genotyping and drug-resistance mutations

The GD HBV phylogenetic typing tool, available at https://www.genomedetective.com/app/typingtool/hbv/, was used to infer genotypes, sub-genotypes, and serotypes of HBV genomes and Geno2pheno software, available at http://hbv.geno2pheno.org/, was used to identify potential drug resistance and antibody escape mutations. Briefly, the HBV phylogenetic typing tool utilizes phylogenetic methods to identify the HBV genotype of a nucleotide sequence. Geno2pheno is an online platform that applies an algorithm that is widely used to predict phenotypic drug resistance mutations in the P open reading frames and antibody escape mutations in the S open reading frames of HBV genomes. The sequences from this study were deposited in GenBank under the accession numbers **<u>PP123755 - PP123892</u>.**

### Statistical analyses

All numerical data were analyzed using RStudio *version 4.1.3* (Posit team (2023). RStudio: Integrated Development Environment for R. Posit Software, PBC, Boston, MA. URL http://www.posit.co/). Baseline characteristics were presented as proportions for categorical data, as means for normally distributed continuous data, or as medians for skewed normally distributed variables. Kruskal Wallis tests (for skewed continuous variables) were used to test for differences in age and viral load groupings. A Chi-square test was used to determine the statistical significance of differences in the gender classes. A Pearson linear correlation test was used to test for a correlation between viral loads and genome coverage of viral sequences and a pairwise t-test was used to test for differences in genome coverage between the different HBV genotypes.

## Authors’ contributions

TdO conceptualized the project, participated in the methodology and investigation, acquired the funding, administered the project, and supervised the project. DPM conceptualized the project, assisted in the methodology, investigation, and visualization, and supervised the project. DT conceptualized the project and assisted in the methodology, investigation, visualization, and writing of the original draft. WC conceptualized the project and assisted in the methodology, investigation, and writing of the original draft. SEJ conceptualized the project, assisted in the methodology investigation, and writing of the original draft. TM conceptualized the project and assisted in the methodology and investigation. WP assisted in the methodology. GvZ assisted in the methodology. JG assisted in the methodology. SP assisted in the methodology. UJA assisted in the methodology. TJS assisted in the methodology. YN assisted in the methodology. All authors read and approved the final manuscript.

## Data Availability

All data produced within the manuscript are available on publically available databases.

## Acknowledgments

This research was supported in part by the South African Medical Research Council (SAMRC) with funds received from the National Department of Health. Sequencing activities at KRISP and CERI are supported in part by grants from the Rockefeller Foundation (HTH 017), the Abbott Pandemic Defence Coalition (APDC), the National Institute of Health USA (U01 AI151698) for the United World Antivirus Research Network (UWARN) and the INFORM Africa project through IHVN (U54 TW012041), the SAMRC South African mRNA Vaccine Consortium (SAMVAC), the South African Department of Science and Innovation (SA DSI) and the SAMRC under the BRICS JAF #2020/049, European Union supported by the Global Health EDCTP3 Joint Undertaking and its members, European Union’s Horizon Europe Research and Innovation Programme (101046041) and the Health Emergency Preparedness and Response Umbrella Program (HEPR Program), managed by the and the World Bank Group (TF0B8412). The content and findings reported herein are the sole deduction, view, and responsibility of the researcher/s and do not reflect the official position and sentiments of the funding agencies.

