## Supplementary Table 1 and 2 for "An Oxford Nanopore Technology-Based Hepatitis B Virus Sequencing Protocol Suitable For Genomic Surveillance Within Clinical Diagnostic Settings" (from another version of this preprint: version 1, 2024-01-20)

**Table S1: NCBI SRA sample accessions.**

| **Sample ID** | **SRA Accession** |
| --- | --- |
| K048718 | SRR27466843 |
| K048719 | SRR27466842 |
| K048720 | SRR27466818 |
| K048721 | SRR27466807 |
| K048722 | SRR27466837 |
| K048723 | SRR27466832 |
| K048724 | SRR27466831 |
| K048725 | SRR27466830 |
| K048726 | SRR27466829 |
| K048727 | SRR27466827 |
| K048728 | SRR27466841 |
| K048729 | SRR27466828 |
| K048730 | SRR27466826 |
| K048731 | SRR27466825 |
| K048732 | SRR27466824 |
| K048734 | SRR27466822 |
| K048735 | SRR27466821 |
| K048736 | SRR27466820 |
| K048739 | SRR27466817 |
| K048740 | SRR27466816 |
| K048741 | SRR27466815 |
| K048743 | SRR27466813 |
| K048744 | SRR27466812 |
| K048746 | SRR27466811 |
| K048747 | SRR27466810 |
| K048749 | SRR27466808 |
| K048750 | SRR27466806 |
| K048751 | SRR27466805 |
| K048752 | SRR27466804 |
| K048753 | SRR27466803 |
| K048754 | SRR27466802 |
| K048756 | SRR27466801 |
| K048757 | SRR27466800 |
| K048760 | SRR27466838 |
| K048762 | SRR27466835 |
| K056885 | SRR26038214 |
| K056886 | SRR26038213 |
| K056887 | SRR26038201 |
| K056888 | SRR26038190 |
| K056889 | SRR26038179 |
| K056890 | SRR26038168 |
| K056891 | SRR26038140 |
| K056892 | SRR26038129 |
| K056893 | SRR26038118 |
| K056894 | SRR26038152 |
| K056895 | SRR26038212 |
| K056896 | SRR26038210 |
| K056897 | SRR26038209 |
| K056898 | SRR26038208 |
| K056899 | SRR26038207 |
| K056900 | SRR26038206 |
| K056901 | SRR26038205 |
| K056902 | SRR26038204 |
| K056903 | SRR26038203 |
| K056904 | SRR26038202 |
| K056905 | SRR26038200 |
| K056906 | SRR26038199 |
| K056907 | SRR26038198 |
| K056908 | SRR26038197 |
| K056909 | SRR26038196 |
| K056910 | SRR26038195 |
| K056911 | SRR26038194 |
| K056912 | SRR26038193 |
| K056913 | SRR26038192 |
| K056914 | SRR26038191 |
| K056915 | SRR26038189 |
| K056916 | SRR26038188 |
| K056917 | SRR26038187 |
| K056918 | SRR26038186 |
| K056919 | SRR26038185 |
| K056920 | SRR26038184 |
| K056921 | SRR26038183 |
| K056922 | SRR26038182 |
| K056923 | SRR26038181 |
| K056924 | SRR26038180 |
| K056925 | SRR26038178 |
| K056926 | SRR26038177 |
| K056927 | SRR26038176 |
| K056928 | SRR26038175 |
| K056929 | SRR26038174 |
| K056930 | SRR26038173 |
| K056931 | SRR26038172 |
| K056932 | SRR26038171 |
| K056933 | SRR26038170 |
| K056934 | SRR26038169 |
| K056935 | SRR26038167 |
| K056936 | SRR26038166 |
| K056937 | SRR26038165 |
| K056938 | SRR26038164 |
| K056939 | SRR26038163 |
| K056940 | SRR26038162 |
| K056941 | SRR26038161 |
| K056942 | SRR26038160 |
| K056943 | SRR26038159 |
| K056944 | SRR26038141 |
| K056945 | SRR26038139 |
| K056946 | SRR26038138 |
| K056947 | SRR26038137 |
| K056948 | SRR26038136 |
| K056949 | SRR26038135 |
| K056950 | SRR26038134 |
| K056951 | SRR26038133 |
| K056952 | SRR26038132 |
| K056953 | SRR26038131 |
| K056954 | SRR26038130 |
| K056955 | SRR26038128 |
| K056956 | SRR26038127 |
| K057252 | SRR26038126 |
| K057253 | SRR26038125 |
| K057254 | SRR26038124 |
| K057255 | SRR26038123 |
| K057256 | SRR26038122 |
| K057257 | SRR26038121 |
| K057258 | SRR26038120 |
| K057259 | SRR26038119 |
| K057260 | SRR26038117 |
| K057261 | SRR26038116 |
| K057262 | SRR26038115 |
| K057263 | SRR26038114 |
| K057264 | SRR26038158 |
| K057265 | SRR26038157 |
| K057266 | SRR26038156 |
| K057267 | SRR26038155 |
| K057268 | SRR26038154 |
| K057269 | SRR26038153 |
| K057270 | SRR26038151 |
| K057271 | SRR26038150 |
| K057272 | SRR26038149 |
| K057273 | SRR26038148 |
| K057274 | SRR26038147 |
| K057275 | SRR26038146 |
| K057276 | SRR26038145 |
| K057277 | SRR26038144 |
| K057278 | SRR26038143 |
| K057279 | SRR26038142 |
| K057280 | SRR26038211 |

**Table S2: Verification of the HBV recombinants by the RDP, GeneConv BootScan, MaxChi, Chimaera, Sister scan, and 3Seq methods implemented in RDP5.46.**

| **Recombinant** | **Minor Parent** | **Major Parent** | **RDP** | **GeneConv** | **BootScan** | **MaxChi** | **Chimaera** | **SisScan** | **3Seq** |
| --- | --- | --- | --- | --- | --- | --- | --- | --- | --- |
| K048723* | A | D | 1.08^-15^ | 5.51^-10^ | 4.61^-02^ | 7.31^-09^ | 1.15^-07^ | 1.58^-04^ | 6.45^-13^ |
| K048737* | D | A | 1.66^-11^ | 3.93^-11^ | NS | 6.41^-07^ | 2.02^-05^ | 1.79^-08^ | 8.29^-10^ |
| K048753* | D | A | 3.09^-10^ | 3.34^-06^ | NS | 5.64^-06^ | 4.17^-05^ | 1.26^-09^ | 8.07^-09^ |
| K048756* | D | A | 1.59^-11^ | 2.21^-08^ | 1.95^-02^ | 1.00^-10^ | 1.00^-10^ | 1.45^-13^ | 1.50^-21^ |
| K056892 | A | A | 2.30^-07^ | 1.76^-05^ | 7.24^-05^ | 2.94^-04^ | 2.01^-04^ | 8.16^-06^ | 9.75^-07^ |
| K056897 | A | AD | 1.89^-21^ | 1.24^-19^ | 1.67^-07^ | 2.60^-12^ | 6.11^-13^ | 1.21^-15^ | 3.58^-25^ |
| K056900* | A | AD | 1.47^-16^ | 8.36^-13^ | 2.77^-13^ | 4.40^-11^ | 4.37^-10^ | 1.64^-13^ | 3.32^-15^ |
| K056911 | D | A | 2.61^-10^ | 3.51^-09^ | 5.81^-05^ | NS | NS | 3.33^-02^ | 1.75^-04^ |
| K056916* | A | D | 2.16^-08^ | 1.92^-08^ | NS | 1.74^-03^ | 1.54^-03^ | 1.14^-04^ | 3.00^-07^ |
| K056934* | AD | A | 7.56^-10^ | 1.39^-05^ | 1.06^-04^ | 3.39^-03^ | NS | NS | 1.77^-04^ |
| K056939 | A | A | 1.86^-06^ | 1.17^-03^ | 7.42^-05^ | 2.05^-05^ | 1.41^-04^ | 3.82^-08^ | 5.02^-04^ |
| K056944* | D | A | 4.52^-18^ | 3.36^-15^ | NS | 1.04^-10^ | 5.47^-11^ | 1.38^-04^ | 1.77^-23^ |
| K056945* | A | AD | 3.42^-20^ | 1.26^-17^ | 1.83^-02^ | 1.88^-18^ | 1.48^-05^ | 2.92^-26^ | 2.53^-27^ |
| K056950* | A | A | 5.16^-08^ | 1.87^-06^ | NS | 7.51^-06^ | 2.34^-06^ | 8.20^-03^ | 4.75^-09^ |
| K057256* | A | D | 1.47^-17^ | 9.40^-17^ | 4.14^-04^ | 4.17^-11^ | 1.31^-11^ | 4.29^-12^ | 4.52^-26^ |
| K057265 |  | A | 7.53^-11^ | 1.72^-09^ | 1.03^-03^ | 4.03^-08^ | 9.05^-05^ | 1.10^-02^ | 1.10^-10^ |
| K057280* | D | A | 6.92^-21^ | 2.63^-22^ | 2.12^-19^ | 3.78^-06^ | 3.29^-06^ | 1.28^-07^ | 8.01^-21^ |

Recombinant IDs marked with a "*" were also classified as recombinants by jpHMM.
